# An intuitive, bimanual, high-throughput QWERTY touch typing neuroprosthesis for people with tetraplegia

**DOI:** 10.1101/2025.04.01.25324990

**Authors:** Justin J. Jude, Hadar Levi-Aharoni, Alexander J. Acosta, Shane B. Allcroft, Claire Nicolas, Bayardo E. Lacayo, Nicholas S. Card, Maitreyee Wairagkar, David M. Brandman, Sergey D. Stavisky, Francis R. Willett, Ziv M. Williams, John D. Simeral, Leigh R. Hochberg, Daniel B. Rubin

## Abstract

Recognizing keyboard typing as a familiar, high information rate communication paradigm, we developed an intracortical brain computer interface (iBCI) typing neuroprosthesis providing bimanual QWERTY keyboard functionality for people with paralysis. Typing with this iBCI involves only attempted finger movements, which are decoded accurately with as few as 30 calibration sentences. Sentence decoding is improved using a 5-gram language model. This typing neuroprosthesis performed well for two iBCI clinical trial participants with tetraplegia - one with ALS and one with spinal cord injury. Typing speed is user-regulated, reaching 110 characters per minute, resulting in 22 words per minute with a word error rate of 1.6%. This resembles able-bodied typing accuracy and provides higher throughput than current state-of-the-art hand motor iBCI decoding. In summary, a typing neuroprosthesis decoding finger movements, provides an intuitive, familiar, and easy-to-learn paradigm for individuals with impaired communication due to paralysis.

---

For individuals with advanced amyotrophic lateral sclerosis (ALS), brainstem stroke, and other conditions that cause paralysis, loss of communication is often cited as one of the most devastating symptoms ^1,2^. People with paralysis affecting both speech and dexterous control of the hands often rely on augmentative and alternative communication (AAC) devices, such as eye-gaze tracking systems, to maintain communication. These devices, however, are often described as being slow, error-prone, and requiring frequent recalibration assisted by care partners, leading to high rates of abandonment ^3,4^.

Intracortical Brain Computer Interfaces (iBCIs) have been used to restore communication for people with paralysis by providing a point-and-click interface ^5–7^, in which a computer cursor can be used to select letters from a virtual on-screen keyboard. Though these systems have the advantage that they can be rapidly calibrated ^8,9^, communication rates are limited by the speed at which the user can point-and-click on individual letters on a computer screen.

More recently, iBCIs have been shown to be effective in enabling higher throughput communication, by means of articulator-based speech decoding ^10–15^ and character-based handwriting decoding ^16^. In these approaches, neural activity pertaining to intended movement, in the absence of actual movement of speech articulators or hand effectors, can be used to decode sequences of phonemes or characters to construct language. High accuracy is maintained with language models ^17^ that take sequences of predicted phonemes or characters and select the most likely intended output sequence based on statistics of large language datasets ^18^. Although iBCI handwriting decoding ^16^ may provide high throughput communication, many individuals may prefer a keyboard-based communication interface due to its well-established integration with digital communication interfaces and their extensive experience with ten-finger typing.

Several recent iBCI studies show accurate neural decoding of 5 ^19^ and 10-dimensional ^20^ finger movements and several hand based gestures ^21–23^, suggesting that higher-dimensional finger movement decoding necessary to enable QWERTY keyboard typing may be feasible. Indeed, recent work ^24^ shows fast and accurate character decoding from neural data pertaining to fine bimanual finger movements, albeit at the single character level. Typing decoding has also recently been performed in able-bodied users with a surface electromyography (sEMG) bracelet ^25^ (though this may be less effective in individuals with severe paralysis due to loss of volitional peripheral nerve activity) and magnetoencephalography (MEG) based recordings ^26^. While both of these recording methods do not require implanted sensors, performance has not yet reached levels required for accurate and reliable communication.

In this work, we introduce a touch typing neuroprosthesis used by two BrainGate2 clinical trial participants to communicate rapidly and accurately. The iBCI typing interface only requires attempted finger movements to decode characters resulting in sentences. The requisite finger flexion and extension intentions closely mimic QWERTY keyboard use in the able-bodied population, taking advantage of familiar finger movement imagery and memory of relative key positions to allow for faster learning of the system. In addition, typing is completely self-paced, and therefore the speed of communication is potentially far greater than AAC devices and previous iBCI typing interfaces that rely on fixed decoding intervals ^24^.

iBCI decoders can have difficulty distinguishing between similar attempted movements; such as similarly shaped characters for handwriting ^16^, similarly articulated phonemes for speech ^10–15^ and nearby finger movements for bimanual typing. However, it may be the case that language models are less sensitive to some patterns of confusion when considering final word-level accuracy. We hypothesize that QWERTY may be more advantageous than handwriting for a given character-level accuracy, as adjacent keys may be less likely to be contiguous or interchangeable with each other in the English language ^27,28^, as opposed to similarly shaped characters.

Taken together, this typing neuroprosthesis has the potential to provide a rapid, highly accurate, and intuitive means of communication for people with paralysis.

## Results

### Neural representation of attempted typing-like finger movements

As an initial assessment, we asked both participants (Fig. 1a,b) to attempt a series of isolated finger movements. In an instructed delay task (Fig. 1c), each participant was presented with a single finger movement cue per trial. Three varying extension and flexion movements (Fig. 1d) were instructed for each finger to ascertain the distinguishability of neural activity pertaining to a total of 30 finger movements required for QWERTY keyboard typing. Participants were instructed to attempt to perform the finger movement cued in text on the screen in front of them when the cue rectangle on screen changed from red to green.

**Figure 1:**
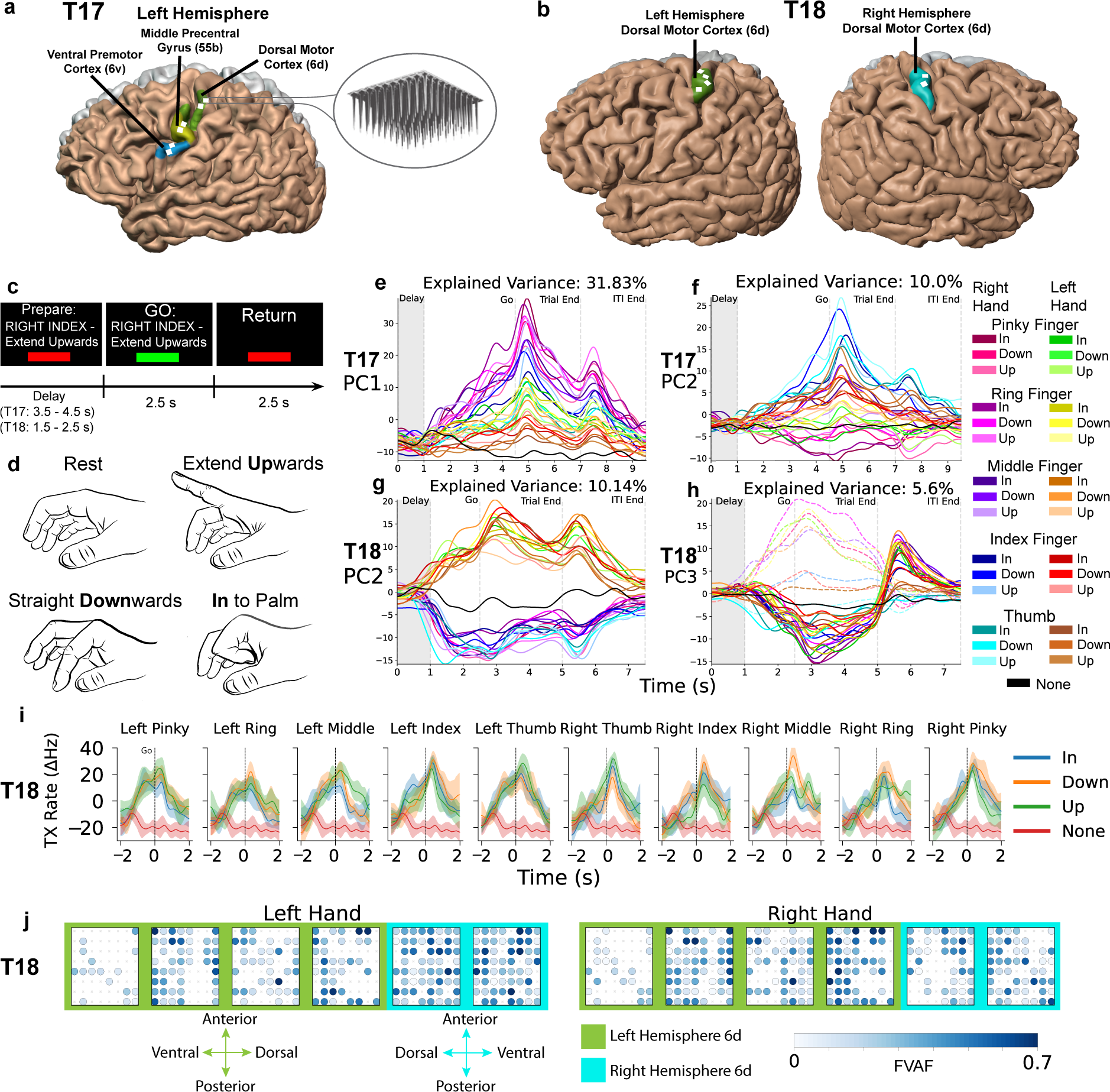
Neural tuning for contralateral and ipsilateral finger movements is present in pre-central gyrus. **a**, T17 microelectrode array locations in the left hemisphere. Two arrays were placed in each of the following cortical areas: dorsal (6d), middle (55b) and ventral (6v) pre-central gyrus. **b**, Left and right hemisphere views of T18 microelectrode array locations. Four arrays were placed in the left, and two arrays in the right dorsal pre-central gyrus (area 6d). **c**, Instructed delay isolated finger movement task. Each participant was instructed to return to rest between trials, and to hold the finger posture throughout the go cue. **d**, Rest position, extend upwards, straight downwards and in to palm movements. **e-h**, Traces are color ordered by hand laterality and secondarily by movement type. Gray shaded regions account for delay variability. **e,f**, The first and second principal components of trial-averaged T17 threshold crossing spike features from the 6v and 6d arrays (256 electrodes) across each condition. **g,h**, The second and third principal components of trial-averaged T18 threshold crossing spike features across each condition from all 384 electrodes. **h**, Extend Upward finger movements on each finger are represented by dashed lines for clarity. **i**, Each panel shows the mean responses of an example electrode in left dorsal premotor cortex from T18 tuned to all flexion and extension positions for every finger. Each line shows mean change in threshold crossing rate averaged across all trials of a given movement condition. Shaded regions indicate a 95% confidence interval. **j**, Tuning heatmaps for all six T18 arrays, backgrounds shaded by hemisphere. Circles indicate that threshold crossing features on a given electrode varied significantly across all 15 unique finger movements on each hand (p < 1 × 10^-5^ assessed with one-way analysis of variance). Shading indicates the fraction of variance accounted for (FVAF) by cross movement differences in threshold crossing rate.

For both participants, there was clear tuning for these discrete finger movements across many electrodes (Extended Data Fig. 3). The trial-averaged multiunit neural activity (change in threshold crossing firing rates) for a well-tuned example electrode in T18 (Fig. 1i), showed prominent responses to each of the three finger movements on all ten fingers. For T17, we observed electrodes in both cortical areas 6d (hand motor) and 6v (speech motor) with responses to almost all finger movements on each finger (Extended Data Fig. 1).

For T18, we found that tuning to finger movements on both hands was intermixed at the single electrode level in left precentral gyrus arrays (Fig. 1j), whereas single electrodes in right precentral gyrus arrays tended to have stronger tuning to finger movements on the left hand. For each hand, there was a higher number of tuned electrodes contralaterally than ipsilaterally (Extended Data Fig. 3). In T17 we observed strong tuning to finger movements on both hands in 6d electrodes, with some electrodes in 6v, particularly in the more dorsal 6v array, exhibiting similar tuning to finger movements on both hands (Extended Data Fig. 2).

To visualize activity at the population level, we performed dimensionality reduction through trial-averaged principal component analysis (PCA) of neural trajectories during trials of the isolated finger movement task (Fig. 1e-h). For both participants we observed anatomically segmented neural trajectories in several principal components, primarily separating based on hand and finger position. Furthermore, we observed distinguishable trajectories across time for unique finger movements, particularly in T17.

For example, the first two principal components (PCs) extracted from 6v and 6d electrodes (a total of 256 electrodes, Fig. 1e,f) showed separation between all 30 finger movements in T17. The first PC for T17 (Fig. 1e) showed hand-dependent activity, with right hand cues evoking larger firing rate modulation over time compared to the left hand. Furthermore, within each hand, more medial fingers (pinky and ring) tend to have higher PC values than more lateral fingers (thumb and index). The second PC for T17 (Fig. 1f) showed overlaid finger ordering in each hand, from thumb to pinky. In T18, the second principal component of trial-averaged PCA extracted from all 384 electrodes (Fig. 1g) shows highly diverging trajectories pertaining to hand-dependent activity. The third PC shows high separability in “Extend Upwards” movements across all ten fingers (shown with dashed lines in Fig. 1h).

### Single-trial decoding performance

Encouraged by separability in the trial-averaged low-dimensional representations between finger movement conditions at the population level (Fig. 1e-h), we next applied a non-linear dimensionality reduction technique to single trials. 2D t-distributed stochastic neighbor embedding (t-SNE) spaces consisting of all individual trials, formed using threshold crossing rates recorded during the go period, reveal a well-separated, low-dimensional space for each participant, with several tight clusters forming for each condition (Fig. 2a,b)

**Figure 2:**
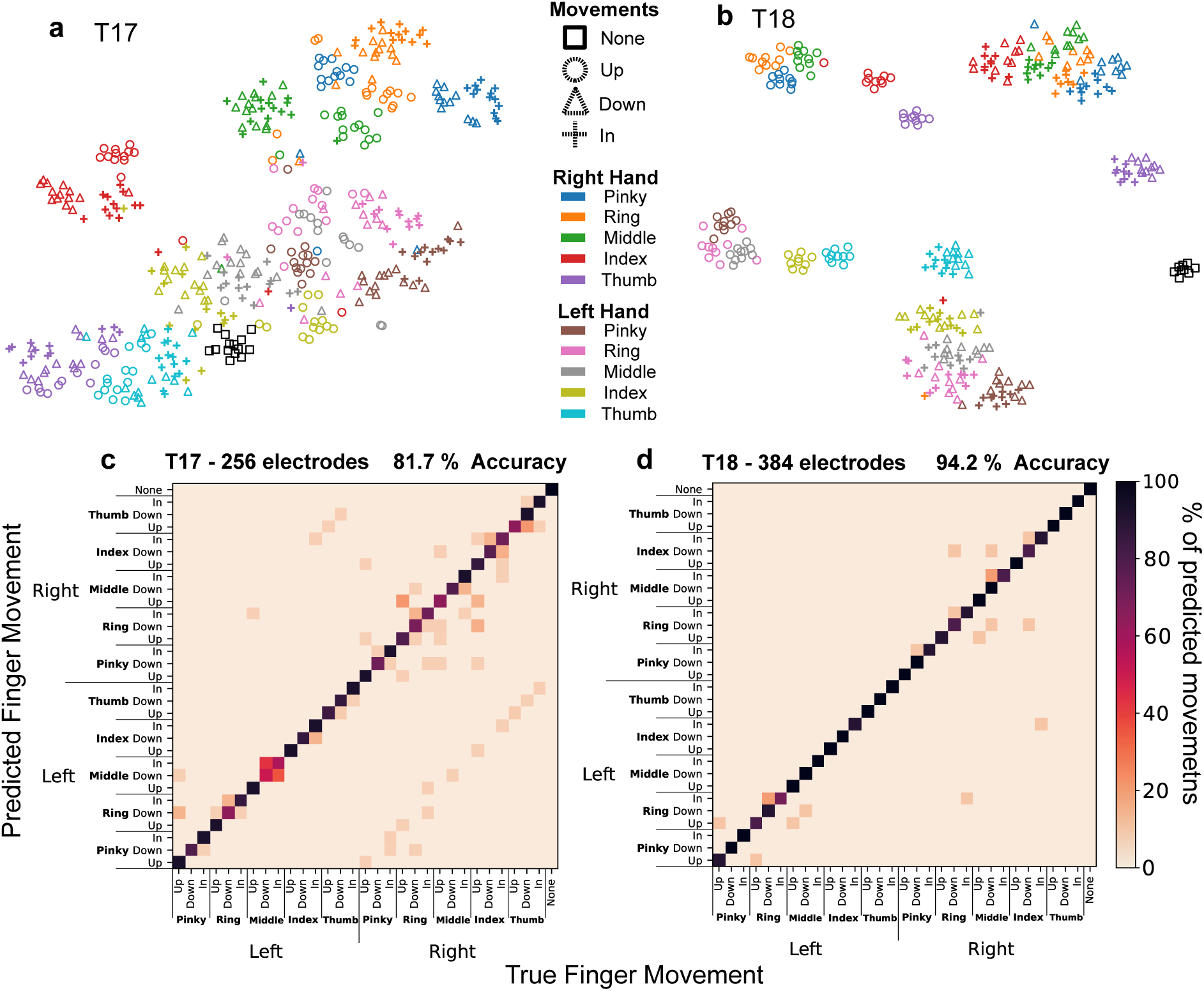
Thirty distinct finger movements can be decoded from PCG. **a,b**, t-distributed stochastic neighbor embedding (t-SNE) low-dimensional representation of all trials of individual finger movements with **a**, T17 and **b**, T18. Fingers are denoted by distinct colors while the three movements within each finger and the no movement condition (None) are denoted by different shapes. **c,d** Confusion matrices showing classification between all thirty isolated finger flexion/extension movements using a Naive Bayes classifier (see Methods) for **c**, T17 and **d**, T18. Colorbar indicates percentage of each true finger movement being decoded as a particular class.

For both participants, trials appeared distinguishable both by finger and by movement type (Fig. 2a,b), with “Extend Upwards” trials (denoted by circles in Fig. 2a,b) generally more distinct than trials of “Straight Downwards” or “In to Palm” on the same finger. This may correspond to the prominently distinct representation of “Extend Upwards” seen in the low-dimensional trajectories discussed above (Fig. 1h) for T18 trials. Particularly clear separation of representations of each hand was evident for T18 (Fig. 2b), which again likely corresponds to the clear divergence of neural activity observed in the low-dimensional state-space representations of neural activity described above (Fig. 1g).

To assess our ability to accurately decode individual finger movements, we trained a Gaussian Naive Bayes (GNB) classifier to distinguish between single trials of each of the thirty discrete movements explored above. For T17 and T18, offline decoding accuracy was 81.7% (95% CI=[79.3, 84.0]) and 94.2% (95% CI=[92.5, 95.8]), respectively (Fig. 2c,d). Somewhat surprisingly, for participant T17 (who only has arrays in left PCG), decoding of left hand movements was more accurate than that of right hand movements. Errors on the left hand were predominantly between movement types within the same finger, whereas on the right hand there was a higher rate of across-finger and across-hand errors (Fig. 2c).

In comparison to T17, T18 exhibited reduced within-hand and within-finger decoding errors (Fig. 2d). This improved performance could be due both the bilateral placement of electrodes as well as the higher number of electrodes in hand motor cortex overall. To understand the relative contribution of bilateral array placement on overall decoding performance, we separately examined ipsilateral and contralateral classification accuracy. Unsurprisingly, for participant T18, unilateral GNB decoding accuracy was lower than bilateral, at 75.4% (95% CI=[72.1, 78.6]) with just left hemisphere arrays and 77.4% (95% CI=[74.4, 80.3]) with just right hemisphere arrays. Unilateral decoding also revealed several cross-hand decoding errors (Extended Data Fig. 4), most notably with left hemisphere arrays. Therefore, bilateral array placement with T18 may reduce across-hand decoding errors, consistent with the representation in Fig. 1g.

### The iBCI bimanual QWERTY keyboard

For the neurally-controlled keyboard, we considered all 26 letters, space (Sp), and punctuation (question mark, comma, period), for a total of 30 unique tokens, arranged according to the QWERTY keyboard layout (Fig. 3a). The keyboard consists of 3 adjacent rows of 10 keys per row. Three movements on each finger, corresponding to the movements performed in the isolated movement task (Fig 1d), allow the user to select any of the 30 keys at any time. Mapped to our keyboard, these movements now correspond to: (1) upward extension from the resting position for an upper row key, (2) straight downwards flexion for a middle row key, and (3) curling into the palm for a bottom row key. Typing speed is controlled by the participant.

**Figure 3:**
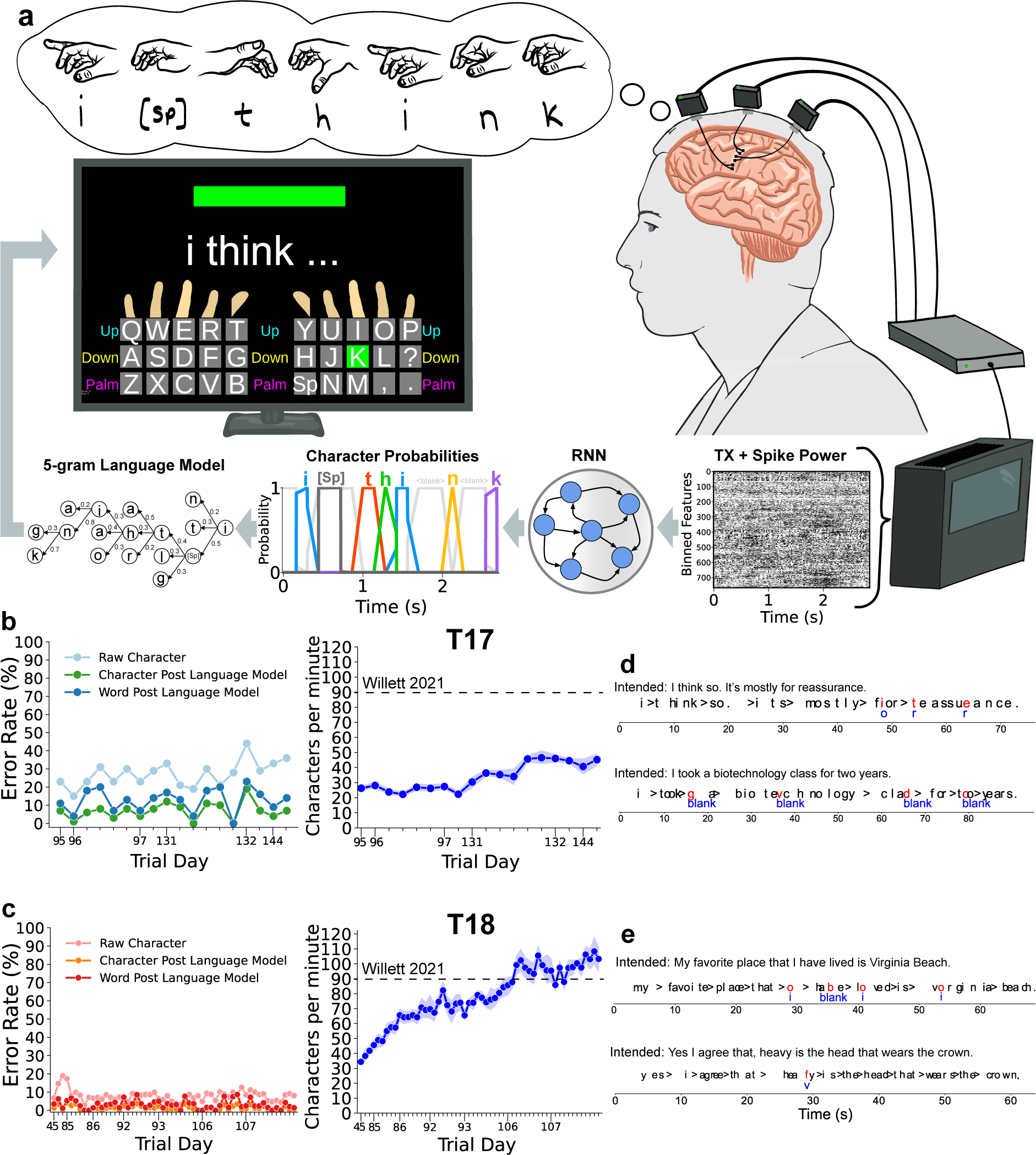
The iBCI bimanual QWERTY keyboard affords high throughput communication. **a**, The keyboard layout closely mimics a QWERTY keyboard. Finger and text guides on the user interface aid in matching finger movements to keyboard characters. Sentences are constructed by the participant performing a series of finger movements corresponding to the keyboard layout shown. Character probabilities are predicted by a recurrent neural network (RNN) from neural features. Character probabilities are continuously considered by the language model, with the currently predicted sentence updated on screen. **b**, T17 and **c**, T18, Left: Mean raw character, character post language model and word post language model error rates shown across all sessions. Numbered x-axis ticks indicate trial days, followed by blocks per day. Each circular marker indicates a block. Right: Mean characters per minute typed by each participant across all sessions per block. Shaded regions indicate a 95% confidence interval. **d**, T17 and **e**, T18 real-time example sentence decoding during free-typing. Spaces are indicated with the greater than sign (’>’). Red characters indicate errors, blue characters below incorrect characters indicate the second most probable character predicted by the RNN.

Character decoding was performed by a recurrent neural network (RNN), which inferred character probabilities in sequence as the user performed a series of finger movements (Fig. 3a). RNN training (see Methods) minimizes a connectionist temporal classification (CTC) ^29–31^ loss function, which aligns binned neural features and sequences of characters. During sentence typing (Fig. 3a), the key corresponding to the most probable character inferred by the RNN is highlighted in green on the virtual keyboard, providing real-time feedback to the user. A 5-gram language model ^17,32,33^ predicts likely sentences given character probabilities inferred by the RNN.

In each session, we evaluated system performance by first training the RNN on calibration blocks of sentences. The RNN was jointly trained with all blocks from all previous sessions along with the current day’s session blocks (see Methods for a listing of trial days). The pipeline outlined above (Fig. 3a) was used to decode sentence typing in real-time, either in a sentence copy task in which the participant was asked to copy a sentence shown on screen or in a free typing task in which the participant could type any sentence they wished. For both the sentence copy and free typing tasks, sentences used for performance evaluation had never been previously seen by the decoder. Sentence origination and conclusion were self-induced in the free typing task and intended sentences were confirmed with the participant.

For T17, mean raw character error rates fluctuated depending on the session, with average word error rates (Fig. 3b) predominantly below 15%. For T18, raw character error rate was consistently low, remaining below 12% after the first trial day. Resulting word error rates for T18 were below 7% across all blocks across all sessions, with several blocks with a word error rate of 0% (Fig. 3c). For both participants, communication rates generally improved over subsequent sessions, with T17 reaching a peak of 47 characters per minute and T18 reaching 110 characters per minute. T18’s speed was faster than the previous state-of-the-art in hand motor iBCI based language decoding using a handwriting strategy ^16^, and approached the iBCI speech-based communication speed of recent work (31.6 words per minute) ^13^ (Extended Data Fig. 6). Encouragingly, this consistent increase over time in typing speed for both participants did not impact character decoding error rate (Extended Data Fig. 5). Neither participant was a touch typist prior to disease or injury onset, and as such, both were acclimatizing to the keyboard layout through typing experience as sessions progressed.

Review of the decoded sentences from the free-typing task highlights the robustness of the pipeline (Fig. 3d,e). In many instances, the sequence of raw character output (i.e., the most probable characters predicted by the RNN) was recognizable, even before the language model step. In other cases, the correct character was the second most probable character predicted by the RNN (highlighted in blue in Fig. 3d,e).

To understand the relative contribution of the language model to word error rate, we examined the pattern of errors produced by the RNN when decoding typing and quantified for each character the frequency of specific RNN substitutions required to produce a correct sentence (Fig. 4a,b). For both T17 and T18, we find that decoding errors were most frequent within the same finger or across adjacent fingers.

**Figure 4:**
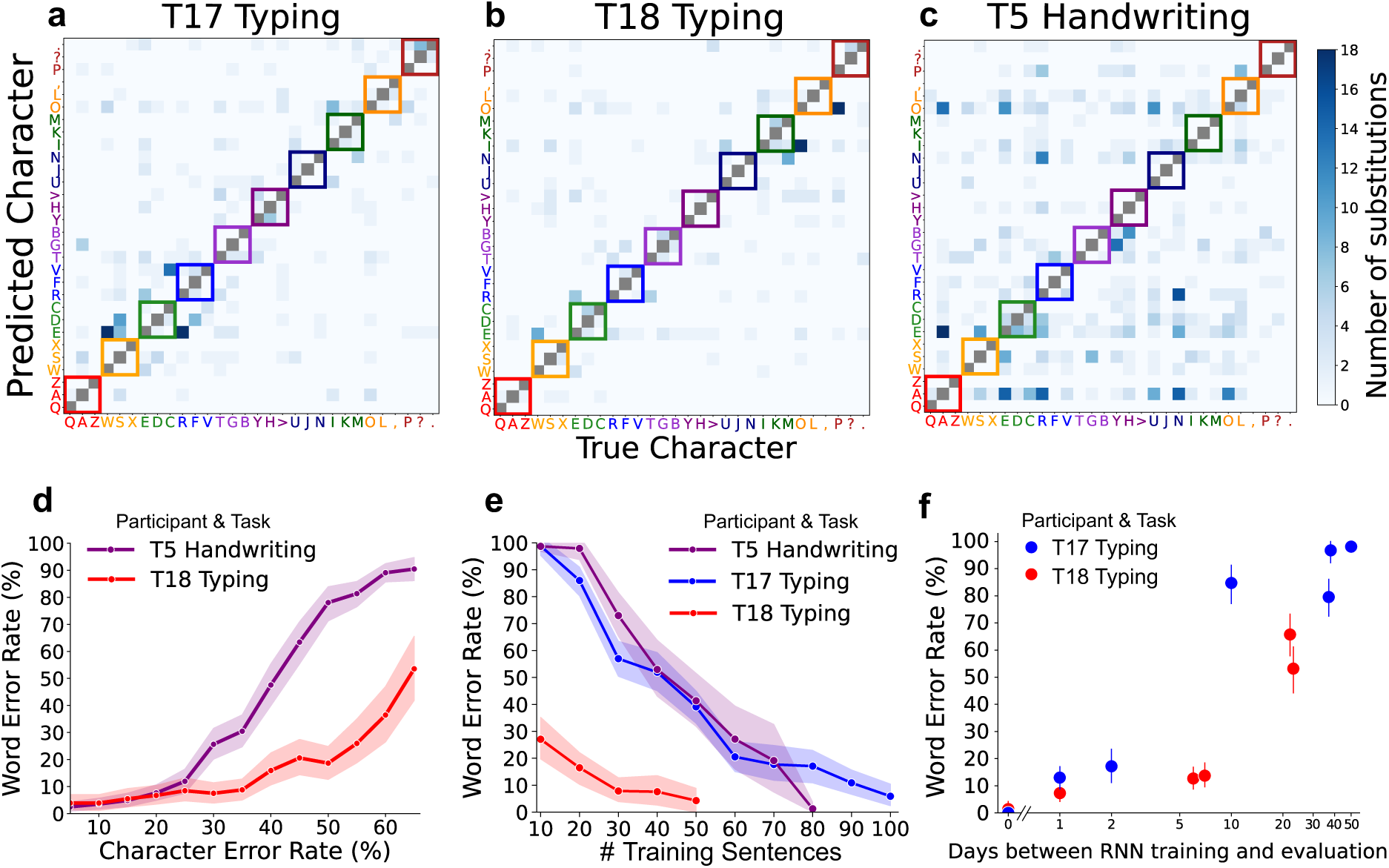
**a,b,c**, Number of substitutions indicates the number of instances of a character on the x-axis requiring substitution with a character on the y-axis in order to generate a correct sentence. Colored boxes indicate characters belonging to the same finger in the typing task, with characters and boxes in darker shades belonging to the right hand (upper diagonal), and lighter shades belonging to the left hand (lower diagonal). **a**, T17 and **b**, T18 offline character substitution RNN decoding errors during the typing task. **c**, T5 ^16^ offline character substitution RNN decoding errors during a handwriting task. Character order and colored boxes belonging to fingers are not relevant to the handwriting task but are shown for comparison. **d**, Mean word error rates resulting from increasing character error rates across participants and tasks. Shaded regions indicate a 95% confidence interval. **e**, Mean word and raw character error rates resulting from an increasing number of RNN training sentences from initialization (no previous RNN training) across participants and tasks. Shaded regions indicate a 95% confidence interval. **f**, Mean word and raw character error rate when training an RNN on day 0 and evaluating decoding performance on held-out sentences whose neural data was recorded on an increasing number of days into the future without RNN recalibration. Bars indicate a 95% confidence interval. X-axis scale is logarithmic.

We compared the effect of these QWERTY error patterns against iBCI handwriting error patterns resulting from offline decoding (using the pipeline in Figure 3a) of a previously published dataset ^16^ (participant T5). In contrast to QWERTY keyboard errors, which were more likely to occur between adjacent keyboard keys, in the handwriting dataset we found that characters that have similar shapes (e.g., {a,e,o,u} and {r,n}) were more likely to be confused (Fig. 4c). This can be detrimental when paired with a language model because these letters are often interchangeable when constructing likely sentences that occur in the English language. A similar phenomenon also occurs in speech decoders ^11–13^, where phonemes which require similar orofacial movements often produce overlapping neural signatures ^12^; these phonemes too (especially vowels) are often interchangeable when a phoneme-based language model considers probable sentences.

We further examined the implication of QWERTY based typing vs. handwriting error patterns on word error rate. Word error rates resulting from increasing character error rates for T18 typing and T5 handwriting reveal that at low character error rates (less than 25%), both participants have similar word error rates; however, at higher character error rates the QWERTY character errors were less likely to result in word errors (Fig. 4d).

We also examined calibration data requirements for simulated novel participants, wherein an RNN was trained with only trials from a single session and evaluated on the character and word decoding accuracy of the remainder of that session’s trials. Notably, with T18, as few as 30 calibration trials were required to obtain a usable 7% word error rate from scratch, approaching 4% at 50 trials (Fig. 4e). With T17, the minimum number of trials required for viable communication was higher, at 100 trials for a 5% word error rate.

To evaluate the stability of the 30-state decoder and look more closely at the rate of change in neural data, we trained an RNN on trials from an early session for both participants and evaluated its decoding performance on future sessions without any recalibration. We found that neural representations in both participants drifted gradually over time (Fig. 4f), with the original decoder for T17 maintaining a word error rate of 15% after 2 days and T18 maintaining a word error rate of 12% after 7 days. The promisingly low word error rate of both participants after 1 day (11% for T17 and 7% for T18) indicates the potential for day-to-day online decoder training on decoded sentences without explicit recalibration ^13,34^.

## Discussion

We introduce a touch typing neuroprosthesis to restore communication for people with paralysis. This system provided the fastest iBCI communication method reported to date based on decoding from hand motor cortex; participant T18 used this system to communicate at 110 characters per minute, which is 20 characters per minute faster than the previous state-of-the-art ^16^. This typing rate of 22 words per minute (WPM) is 81% the speed of able-bodied smartphone typing of 27.3 WPM for T18’s age group ^35^. High speed is achieved while maintaining comparable accuracy to state-of-the-art iBCI character-based decoding ^16,24^. In T17, we demonstrate faster iBCI communication with an unrestricted vocabulary in a person with longstanding anarthria and incomplete locked-in syndrome than previous methods ^36^.

The QWERTY iBCI keyboard employs familiar intended movements, representing an intuitive method of communication. Although less common keyboard layouts such as DVORAK may improve typing speed ^37^ and optimized keyboard layouts may improve resulting word level accuracy ^38,39^, future iBCI users with touch typing experience prior to injury or disease may experience a faster rate of acclimation to the ubiquitous QWERTY keyboard layout, reducing overall time to rapid communication. Once learned, the system may also be used without visual acuity or direct gaze at the screen; as opposed to cursor-based character selection ^5^ which requires knowledge of current cursor position.

Differences between the two participants, including disease and injury etiologies, number and location of recording sites, and other variables may have influenced these initial results. In particular, T18 attained higher online typing speed and accuracy than T17. All six of T18’s Utah arrays were placed in area 6d (containing the anatomical “hand knob” of cortex ^40^), on both the ipsilateral and contralateral hemispheres, for a total of 384 hand motor electrodes (Fig. 1b) – the most reported that we are aware of in the literature. In contrast, T17 has only two arrays in hand motor areas for a total of 128 presumptive hand motor electrodes (Fig. 1a). Despite this unilateral representation, we were able to decode the intended movements of both hands. This is consistent with recent studies demonstrating bilateral representation of intended movement within left motor cortex ^41–45^, and it is reassuring that this decoding approach can be employed by an iBCI user with unilaterally placed recording electrodes. Indeed, decoding solely unilaterally demonstrated redundant bi-hemispheric representation of bimanual finger movements in T18 (Extended Data Fig. 7).

Future work will focus on integrating the typing neuroprosthesis into communication systems to support people with paralysis, in which typed sentences are either vocalized through a text-to-speech (TTS) system or kept on the screen for interaction with a digital communication interface (i.e., email, texting). The use of the language model will be made toggleable as its inclusion does not accommodate arbitrary sequences of characters that do not conform to character sequences often seen in English (e.g., two-factor authentication passwords).

Additional future work will focus on developing independent participant use through online training of the RNN decoder ^13,34^, whereby sentences confirmed to be decoded correctly would be trained upon. This has been shown to be feasible in these data without sentence copy task-based recalibration, due to low word error rates across adjacent days with a frozen decoder (Fig. 4f).

## Data Availability

Data will be made available upon publication.

## Methods

### Clinical Trial

Permission for this study was granted by the U.S. Food and Drug Administration and the Institutional Review Boards of Massachusetts General Hospital, Brown University, and the VA Providence Healthcare System. Research sessions were conducted with two participants, T17 and T18, and data used which was collected previously from a third participant, T5, all of whom were enrolled in the BrainGate2 clinical trial (ClinicalTrials.gov ID: NCT00912041). All research sessions were performed at each participant’s place of residence. This manuscript does not report any clinical trial-related outcomes; instead, it describes scientific and engineering discoveries that were made using data collected in the context of the ongoing clinical trial.

### Participant T17

T17 is a right-handed man in his 30s with ALS. He has tetraplegia, anarthria, and ventilator dependence; his only remaining volitional motor control is over his extraocular muscles. Following enrollment, T17 had a preoperative structural MRI, resting state fMRI, and task-based fMRI to identify appropriate anatomical targets for microelectrode array (MEA) placement. Resting state fMRI was used to generate estimated parcellations of the relevant brain areas using a custom instantiation of the Human Connectome Project ^1,2^ analysis pipeline that was modified for deployment on clinical MRI scanners able to accommodate a mechanical ventilator. Subsequently, T17 underwent placement of six 64-electrode microelectrode arrays (Blackrock Microsystems; 1.5 mm electrode length) in the left precentral gyrus; two arrays were placed in the dorsal precentral gyrus (area 6d), two arrays were placed in the ventral precentral gyrus (area 6v), and two arrays were placed in middle precentral gyrus (area 55b) (Fig. 1a). T17 consented to publication of photographs and videos containing his likeness.

### Participant T18

T18 is a right-handed man in his 40s with tetraplegia resulting from a cervical spinal cord injury (C4 ASIA A) that occurred approximately 18 months prior to trial enrollment. He had a preoperative structural MRI, task-based functional MRI, and resting state fMRI to allow for functional parcellation using the Human Connectome Project (HCP) ^1,2^ pipeline to identify putative targets for microelectrode array placement. He subsequently had four 64-electrode microelectrode arrays (Blackrock Microsystems; 1.5 mm electrode length) placed in the dorsal precentral gyrus (area 6d) of the left (motor dominant) hemisphere and two 64-electrode microelectrode arrays placed in the dorsal precentral gyrus (area 6d) of the right hemisphere (Fig. 1b). T18 consented to publication of photographs and videos containing his likeness.

### Participant T5

T5 is a right-handed man, in his 60s at the time of data collection, with a C4 AIS C (ASIA Impairment Scale C – Motor Incomplete) spinal cord injury that occurred approximately 9 years prior to study enrollment. Two 96-electrode microelectrode arrays (Blackrock Microsystems; 1.5-mm electrode length), were placed in the hand “knob” area of T5’s left-hemisphere (dominant) precentral gyrus. Data are reported from post-implant days 994 to 1246. T5 handwriting data is publicly available at https://datadryad.org/stash/dataset/doi:10.5061/dryad.wh70rxwmv.

### Neural Signal Processing

Neural voltage time series signals were recorded and digitized (30 kHz, 16 bits per sample) using the Neuroplex-E system (Blackrock Neurotech) attached to three percutaneous connectors on each participant’s head, and transmitted via three mini-HMDI cables (one to each Neuroplex headstage), attached to two Gemini hubs (Blackrock Neurotech), prior to final processing via a Neural Signal Processor (NSP) (Blackrock Neurotech). Packets of neural data were streamed from the NSP to our processing pipeline. Signals were analog filtered (4th order Butterworth with corners at 250 Hz to 5 kHz) using the Scipy python library (scipy.signal.filtfilt).

Linear regression referencing ^3^ (LRR) filter coefficients and subsequent electrode-specific thresholds were determined using the filtered digitized data recorded in an initial reference block at the beginning of each session. LRR coefficients are computed by solving *Y* = *WX* where *Y* is the signal from a given electrode we require and *X* is the signal from all other electrodes. We solve for the LRR weight matrix through least squares calculation *W* : *W* = *inv*(*X^T^ X*)*X^T^ Y* where *inv* is matrix inversion ^4^. Electrode-specific thresholds were calculated using filtered 30kHz data once these calculated references had been applied to identify spike events. Thresholds were set at -3.5 times the standard deviation of the voltage signal per electrode. The number of non-causal threshold crossings ^5^ (ncTX) were computed by counting the number of times the filtered neural time series crossed these calculated thresholds (threshold crossing rates). Additionally, spike band power was computed by taking the sum of squared voltages observed during each 10ms time bin. During closed-loop decoding blocks, feature normalization was employed to account for neural nonstationarities (drifts in mean firing rate) which could arise over the course of a block. Within each electrode, threshold crossing rates and spike band power were z-scored (mean subtracted and divided by standard deviation per electrode). Feature extraction (threshold crossings and spike band power), binning, decoding and task phase control were performed through the Python based, modular BRAND ^6^ framework, where each process is instantiated as a self-contained node-based Python program. Messaging between these nodes is performed using a Redis database.

### Isolated Finger Movements

In this instructed delay paradigm, each trial’s preparatory time varied between 3.5 and 4.5 seconds for T17 (due to the participant’s preference to accommodate poorer baseline visual acuity) and between 1.5 and 2.5 seconds for T18. Both participants had a 2.5 seconds movement time (including a hold), and 2.5 seconds rest time. Each participant was instructed to attempt to maintain the rest position (shown in Fig. 1d) until the go cue (rectangle on screen turns green accompanied by a beep tone), whereupon they were instructed to attempt and hold one of the three active movements shown in Fig. 1d for a given finger until “Return” is displayed on screen, accompanied by a click sound. Additionally, a “Do Nothing” condition instructed the participant to remain in the rest position and perform no (intended) movement throughout the Go period.

### Principal Component Analysis (PCA) trajectories

To visualize the neural geometry of thirty isolated finger movements, we plotted the first and second (T17) and second and third (T18) principal components by Gaussian smoothing (standard deviation of 5) all trials over time, then averaging all trials pertaining to each movement condition. In the case of T18 (where each trial was 7500ms in total), a 3D matrix of shape (31, 750, 384) was formed where 31 is the total number of isolated movement conditions, 750 is the number of 10ms time bins, and 384 is the number of threshold crossing features used. The time dimension was preserved and stacked in order to form a 2D matrix of shape (31 * 750, 384) before applying the Scikit-learn ^7^ implementation of PCA, which after reshaping, produces a 3D matrix of shape (31, 750, 3). We then plot each of 31 conditions over 750 time bins for the second and third principal components. Averaged PCA trajectories were Gaussian smoothed with a kernel standard deviation of 20.

### Peristimulus time histograms (PSTHs)

To visualize individual neuronal responses to stimuli, we demonstrate firing rate changes for a representative neuron in T18 upon presentation of the go cue. Threshold crossing rates for each condition for each finger (Up, Down, and In) were mean averaged and Gaussian smoothed (standard deviation of 8) across the time axis. The resulting time series shown in Figure 1i depict the average threshold crossing rates across time for each stimulus (finger movement condition).

### t-distributed stochastic neighbor embedding (t-SNE)

t-SNE is a nonlinear dimensionality reduction technique which preserves local pairwise similarities between samples. We visualize the high-dimensional neural data pertaining to isolated finger movements via the low-dimensional trial by trial similarity space rendered using t-SNE, which is performed on time averaged neural activity across all trials. Time averaging produces a 2D matrix of shape (number of trials, 384), where 384 is the number of threshold crossing features. We use the Scikit-learn ^7^ implementation of t-SNE with a perplexity of 10 (favoring local versus global structure) and PCA initialization.

### Gaussian Naive Bayes

The Naive Bayes classifier is a simple probabilistic predictor which assumes conditional independence between features given the class. Gaussian Naive Bayes takes this a step further for continuous data and makes the additional assumption that values associated with each class are normally distributed. Offline single-trial finger movement classification results (reported in Fig 2c,d) were generated using a cross-validated (leave-one-out) Gaussian Naive Bayes classifier. All trials are z-scored (mean subtracted and standard deviation divided) and time-averaged in the range 80ms before the go cue to 200ms after the go cue for both participants. For every trial, the classifier is trained on all other trials and evaluated on the held-out trial. We use the Scikit-learn ^7^ implementation of Gaussian Naive Bayes.

### Recurrent Neural Network

This decoding pipeline has its origins in automatic speech recognition (ASR) research ^8^, and is utilized in several other recent works, particularly iBCI and electrocorticography (ECoG) based speech decoding ^9–11^. The recurrent neural network (RNN) used for character sequence decoding is implemented in Tensorflow2 ^12^ as a 5 layer gated recurrent unit (GRU) network, each with 512 units. Training using backpropagation through time (BPTT) minimizes the Connectionist Temporal Classification loss function ^13–15^, which aligns binned neural features and sequences of characters. Various regularization and data augmentation techniques are utilized: Dropout, Gaussian White noise, L2 weight norm. Hyperparameter details can be found in Supplemental Table 1. For T17, electrodes from areas 6v and 6d are used for sentence typing decoding, for a total of 256 electrodes (electrodes from area 55b are omitted). For T18, all electrodes are used, for a total of 384 electrodes. RNN inference is performed using threshold crossing and spike band power features for a total of 768 features for T18 and 512 features for T17, which are extracted from raw neural data in real time. These features have been binned into 20ms bins. In the case of T17, character probabilities are emitted by the RNN every 180ms on a preceding window of 320ms of neural features. For T18, character probabilities are inferred every 120ms on a preceding window of 300ms of neural features, owing to the faster typing speed of this participant (see RNN hyperparameters in Supplemental Table 1).

Much recent work focuses on reducing or eliminating the adverse decoding effects of neural nonstationarities ^16–21^. Here, we utilized training data across multiple days; a non-linear input layer is added per session to account for cross-session neural variability, as was used in recent work ^9,11,16^. Each non-linear layer contains the same number of units as the dimensionality of the neural features (768 for T18, 512 for T17). During training, batches of trials only from a given session are selected at random, such that the corresponding input layer is trained along with the 5 layer RNN. This resulted in a cross-session ensemble decoder that was relatively robust to nonstationarities in the short to medium term (resulting in the reported online error rates in Fig. 3b,c)

### WFST Language Model

We use a 5-gram language model implemented using a weighted finite state transducer (WFST), built on the Kaldi system ^22^. This probabilistic model was trained using the WeNet framework ^23^ and utilized the large OpenWebText2 ^24^ corpus to compute conditional probabilities. When given lattices of RNN output probability vectors, the WeNet framework initiates several Viterbi ^25,26^ searches through the most recent lattice, incorporating character-by-character transition probabilities when inferring the most likely typed sentence. Language model parameter details can be found in Supplemental Table 2. Punctuation not contained in the keyboard, such as apostrophes, are inferred by the language model based on decoded sentence context. As the user typed a given sentence, the entire sentence history of character probabilities inferred by the RNN thus far was used by the 5-gram language model ^22,23^ to ascertain the most likely sentence typed given the series of RNN probabilities and sequences of characters likely seen in sequence in English. The most likely sentence deduced by the language model based on RNN output up to that time was continuously displayed on screen.

### Character and Word Error Rate Definitions

Raw character error rate is calculated as the normalized number of character insertions, deletions and substitutions required to produce a correct sentence based on the top-1 RNN predictions at each timestep. Similarly, character error rate post language model is calculated on language model output. Word error rate is calculated as the number of word insertions, deletions and substitutions required to produce a correct sentence, based on language model output. In each case, the total number of errors is normalized by the number of characters or words in a given sentence.

### Free Typing

This conversational mode of typing is not cued, instead the RNN decoder begins decoding as soon as the block commences. As a component of the Connectionist Temporal Classification (CTC) loss function used for RNN training, a blank class is used (in addition to the other 30 keyboard key classes) to account for windows of neural features which do not fit well to any of the 30 keys, and instead represent momentary rest periods between key presses. Thus, while the participant is not typing actively, this blank class is predicted by the RNN. During free typing, this corresponds to no output on screen. Once a non blank class is predicted by the RNN, sentence decoding proceeds as with the sentence copy task, with RNN output probabilities sent to the language model before the most likely typed sentence is output on screen. The end of a sentence is signaled either with the prediction of a period (’.’), a question mark (’?’) or four consecutive seconds of blank class decodes.

### Sentence Evaluation

Ground truth calibration sentences in the sentence copy task were sourced from transcriptions of the Switchboard Corpus ^27^, chosen due to their conversational nature. The number of evaluation sentences in a given block varied from between 10-20 sentences in the sentence copy task to between 4-20 in the free typing task.

### Handwriting vs. Typing substitution errors

Training of the RNN in each instance is stopped early once a character error rate (top-1) of 30% is reached in the confusion matrices in Figures 4a,b and c. Evaluation is then performed on 100 held-out trials from a single session in the case of T17 and T18 and 60 trials from a single session in the case of T5. In the case of Figure 4d, The RNN is trained on data from all sessions for each task and participant, with early stopping of RNN training at each of the indicated character error rates on the x-axis. Evaluation is again performed on 100 held-out trials from a single session in the case of T17 and T18 and 60 trials from a single session in the case of T5.

## Acknowledgments

We thank T17, T18, T5, their families and carepartners for the time and effort they contributed to this research. We thank Dr. Stephen Mernoff for clinical monitoring of trial participants. We thank Dr. Gladys Hill for her illustrations in Figures 1 and 3. We thank Maryam Masood, Dave Rosler, and Beth Travers for their regulatory and team management efforts supporting this work.

This work was supported by:

Office of Research and Development, Department of Veterans Affairs (A2295R, A4820R, N2864C, A3803R), NIH NIDCD (U01DC017844, K23DC021297, R01DC014034), NIH NINDS (U01NS123101), AHA (23SCE-FIA1156586), CDMRP (HT94252310153), a Pilot Award from the Simons Collaboration for the Global Brain (872146SPI), A.P. Giannini Postdoctoral Fellowship, a Career Award at the Scientific Interface from BWF.

Several analyses were performed using code from: https://github.com/fwillett/speechBCI and T5 handwriting data was acquired from https://datadryad.org/stash/dataset/doi:10.5061/dryad.wh70rxwmv.

## Author Contributions

JJJ conceived the study, built the real-time typing decoder, wrote the manuscript, and led the development, analysis, and interpretation of all experiments.

HLA and DBR reviewed analysis and interpretation of experimental data.

FRW designed the real-time sequence decoder paradigm, originally incorporated the n-gram language model into sequence decoding and produced offline analysis tools.

NSC, MW, DMB, and SDS built the real-time BRAND feature extraction system, implemented the neural brain-to-text decoding pipeline in BRAND, built the task control interface, and offline analysis tools.

SBA, MW and NSC wrote software for Neural Signal Processing.

SBA, JDS and JJJ installed hardware for neural recording with T17 and T18.

JJJ and BEL collected T18 typing session data, AJA, HLA, CN and JJJ collected T17 typing session data.

FRW collected T5 handwriting session data.

JDS managed integration and deployment of BCI system software and hardware.

LRH is the sponsor-investigator of the multisite BrainGate2 pilot clinical trial.

ZMW, DBR and LRH planned T17 and T18’s array placement surgeries, and ZMW performed T17 and T18’s array placement surgeries. LRH was responsible for all clinical trial related activity at VA Providence and MGH. LRH and DBR supervised and guided all research activity with T17 and T18.

The study was supervised and guided by DBR.

All authors reviewed and edited the manuscript.

## Extended Data

**Extended Data Figure 1:**
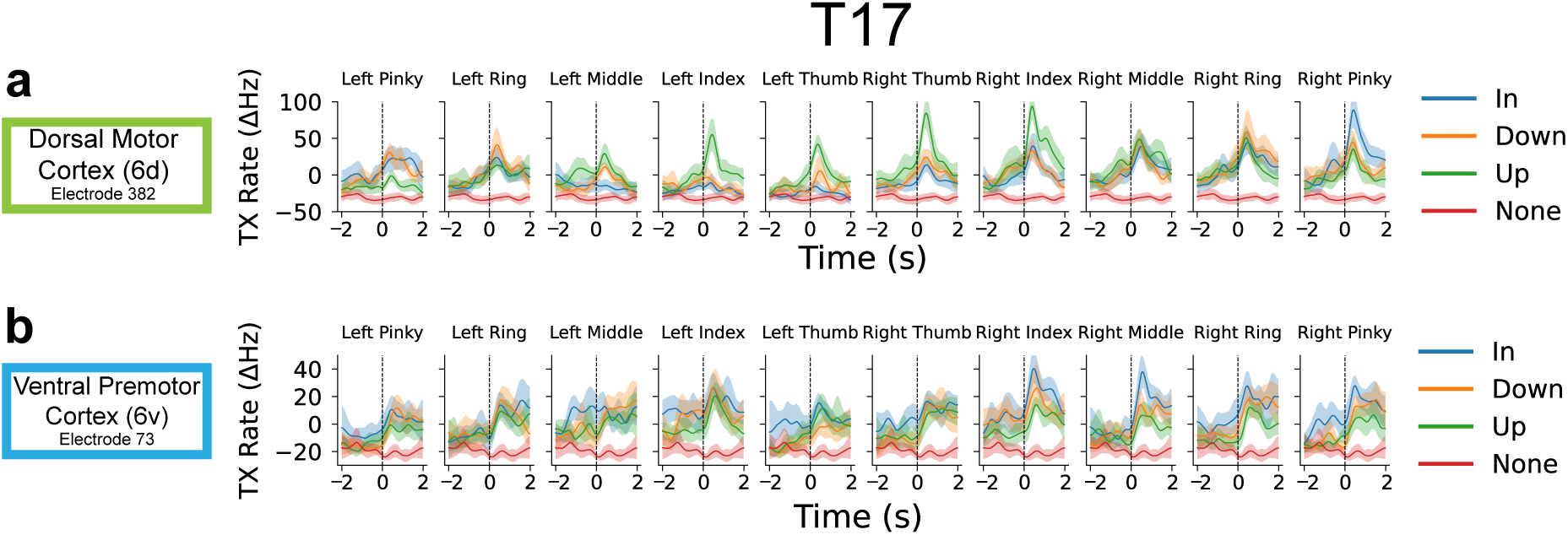
**a**, Each panel shows the mean responses of an example electrode in area 6d of motor cortex (electrode 382, placement in left dorsal motor cortex) from T17 tuned to flexion and extension positions for every finger, where each line shows mean change in threshold crossing rate averaged across all trials of a given movement condition. **b**, Each panel shows the mean responses of an example electrode in speech motor cortex (electrode 73, placement in left ventral premotor cortex, area 6v) from T17, weakly tuned to flexion and extension positions for every finger, where each line shows mean change in threshold crossing rate averaged across all trials of a given movement condition. Shaded regions indicate a 95% confidence interval.

**Extended Data Figure 2:**
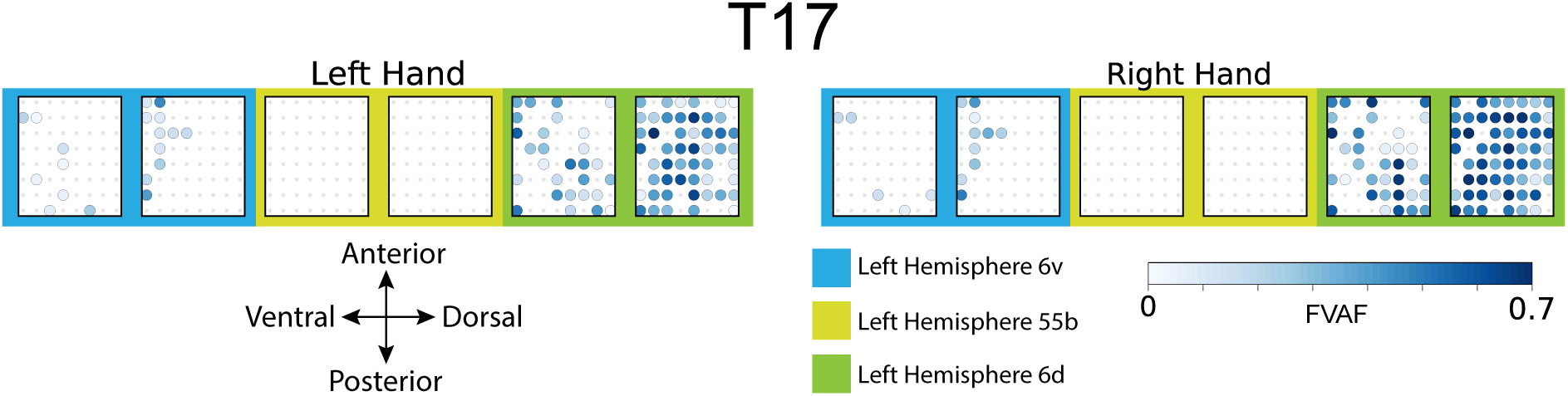
Tuning heatmaps for all 6 T17 arrays, array backgrounds shaded by brain area implanted (all in the left hemisphere). Circles indicate that threshold crossing features on a given electrode varied significantly across all 15 unique finger movements on each hand (p < 1 × 10^-5^ assessed with one-way analysis of variance). Shading indicates the fraction of variance accounted for (FVAF) by cross movement differences in threshold crossing firing rate.

**Extended Data Figure 3:**
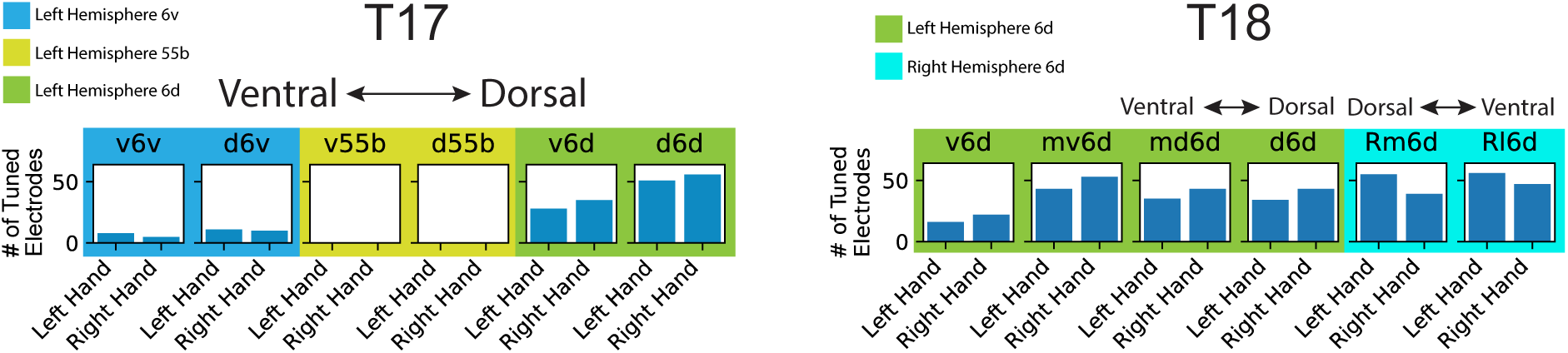
Number of strongly tuned electrodes where threshold crossing features on a given electrode varied significantly across all 15 unique finger movements on each hand (p < 1 × 10^-5^ assessed with one-way analysis of variance) for each participant.

**Extended Data Figure 4:**
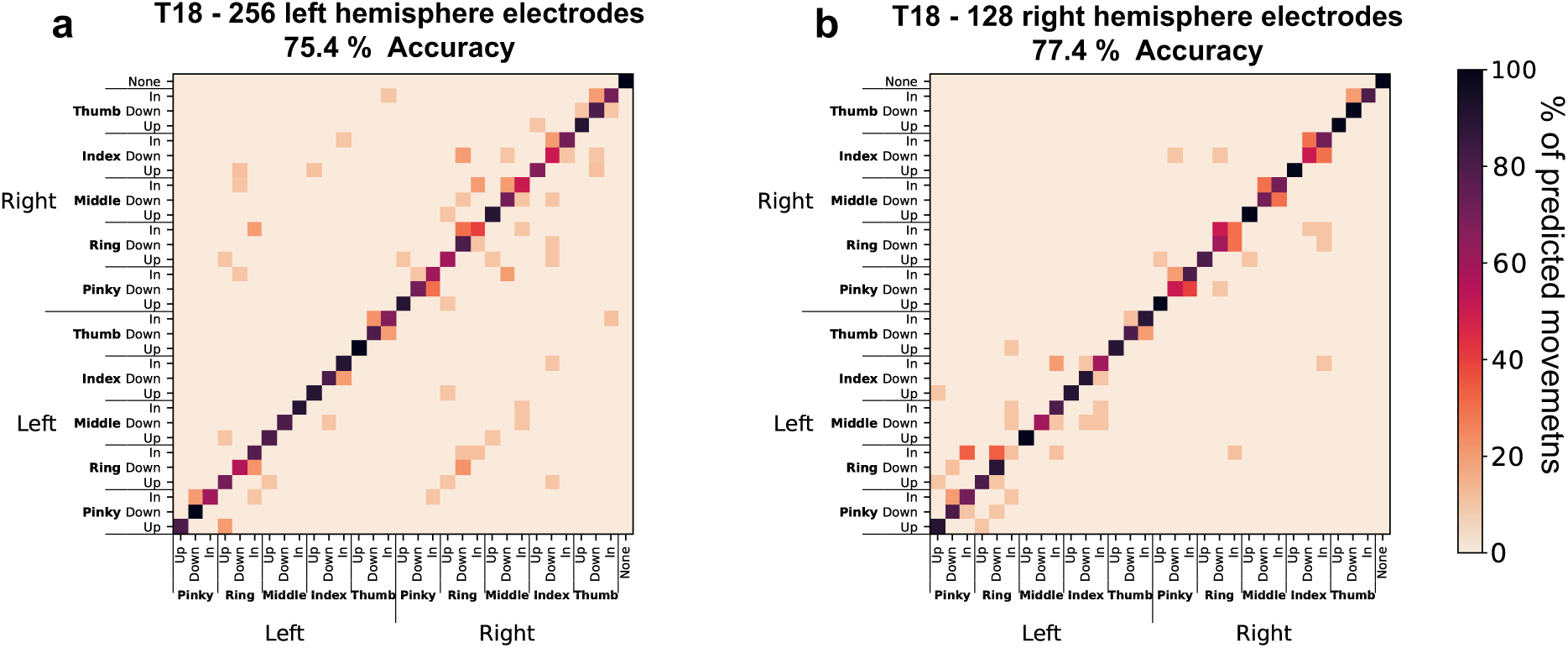
Classification using a Gaussian Naive Bayes (GNB) classifier for T18 with **a**, 256 electrodes placed in left precentral gyrus, resulting in a decoding accuracy of 75.4% (95% CI=[72.1, 78.6]), and **b**, 128 electrodes placed in right precentral gyrus, resulting in a decoding accuracy of 77.4% (95% CI=[74.4, 80.3]).

**Extended Data Figure 5:**
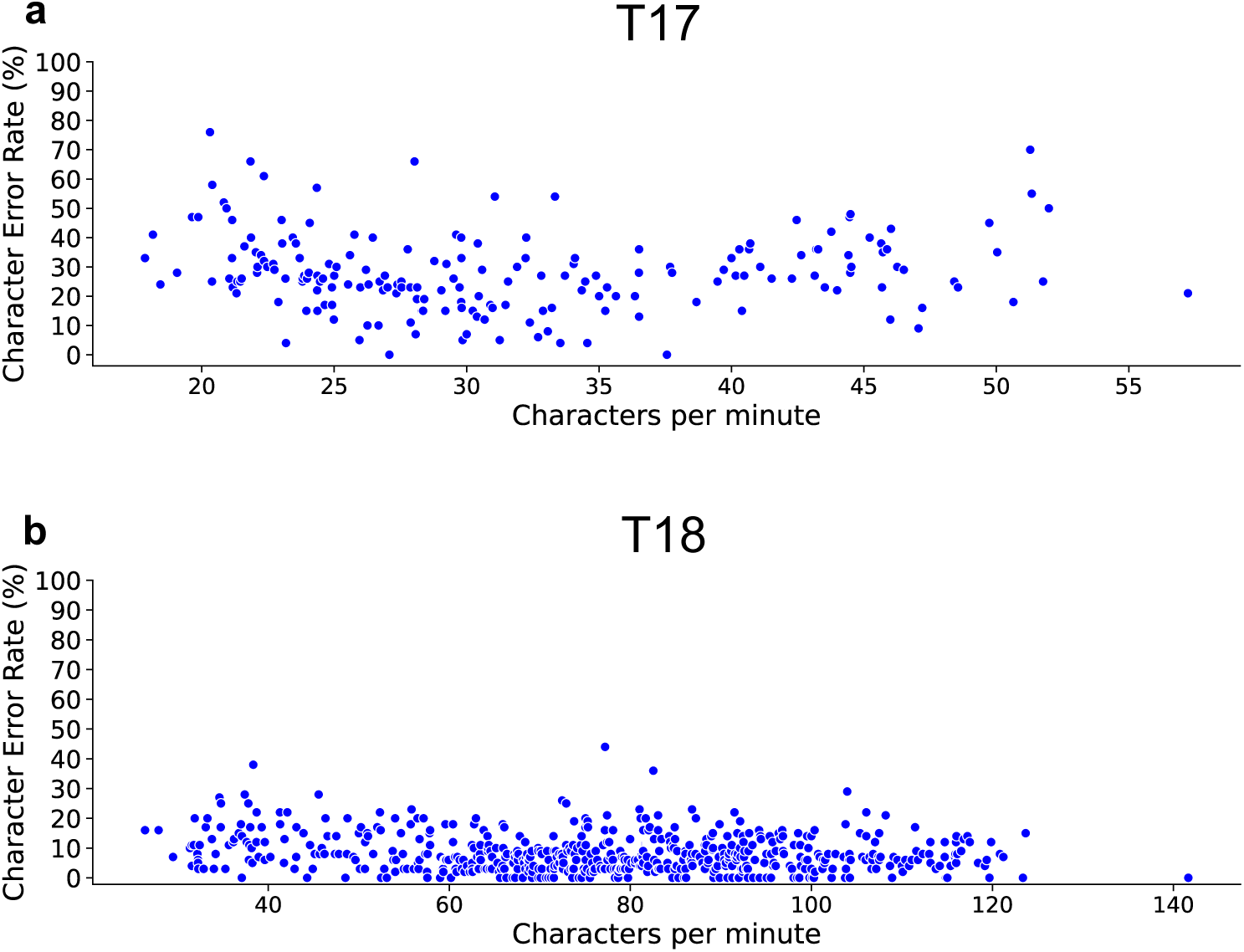
**a,b**, T17 and T18 character error rate resulting from increasing typing speed for all trials. Each circle marker represents a single sentence trial.

**Extended Data Figure 6:**
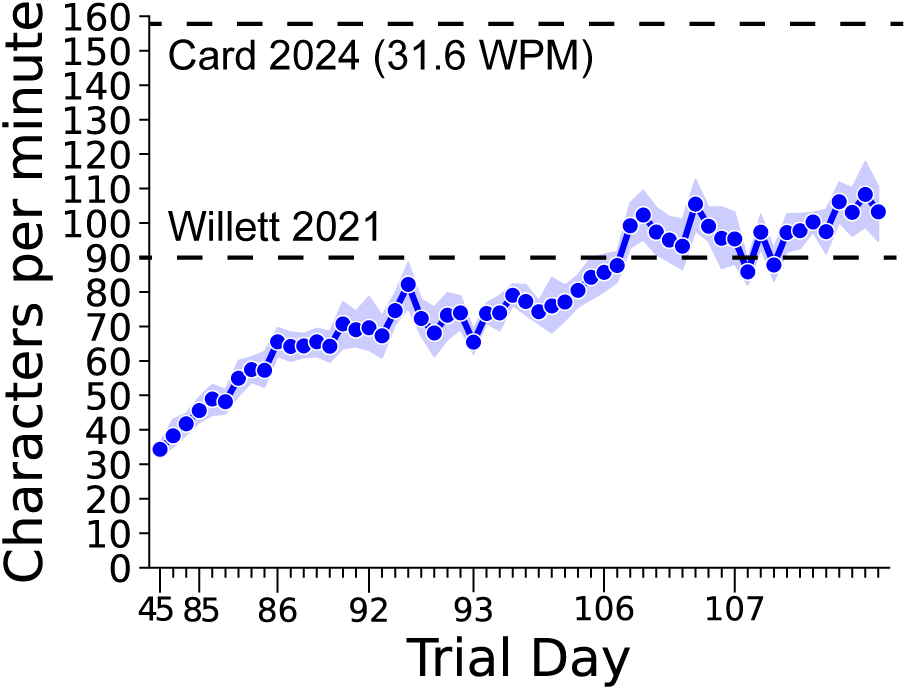
T15 rate of communication as reported in ^11^ was 31.6 words per minute, while T18 typing peaks at an average of 22 words per minute. Thus, our typing neuroprosthesis has a peak communication rate 70% that of T15 attempted speech.

**Extended Data Figure 7:**
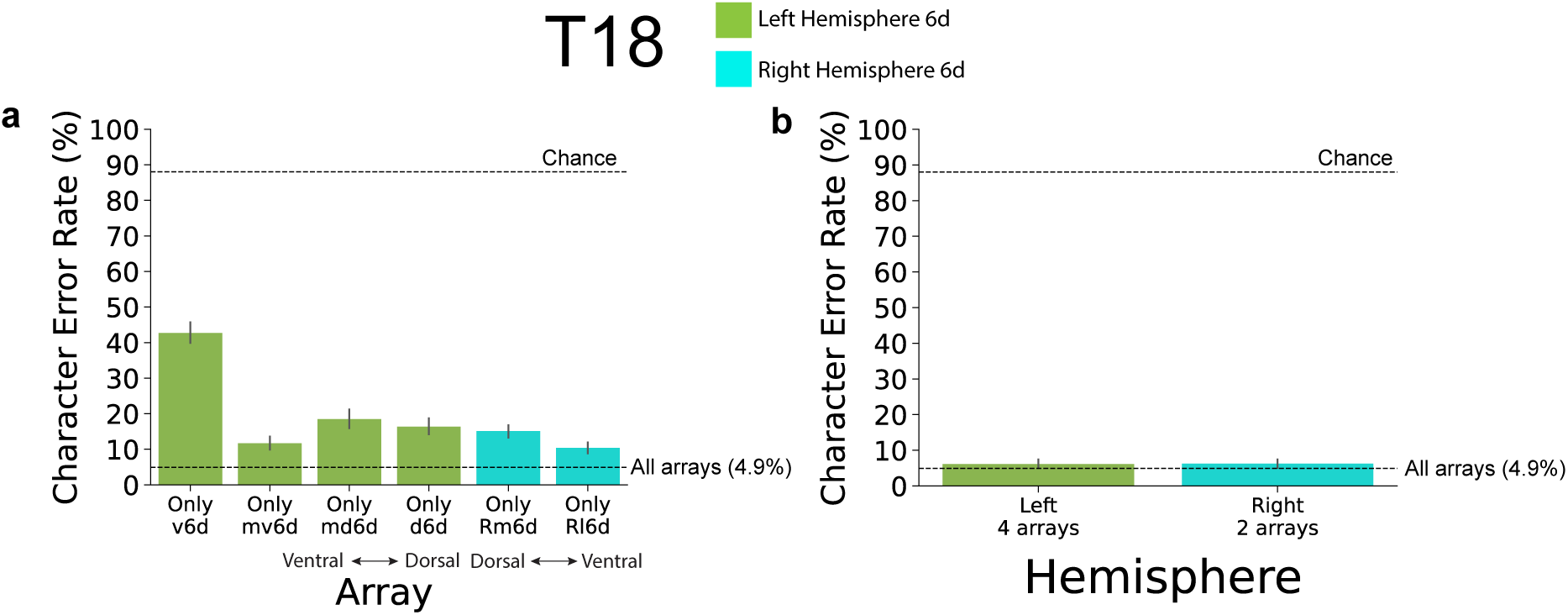
**a**, RNN trained and evaluated offline using each T18 array individually. **b**, RNN trained and evaluated offline using arrays on each T18 hemisphere individually. Chance level character error rate results from always predicting the character “e”.

## Supplement

**Supplemental Table 1:** RNN decoding model parameters.

| Parameter Name | Parameter Description | Value |
| --- | --- | --- |
| # Units | Units in each GRU layer | 512 |
| # Layers | Number of GRU layers | 5 |
| Kernel Size | Number of feature time bins input to the RNN model (size of sliding window) | T17: 16<br>T18: 15<br>T5: 24 |
| Kernel Stride | Size of sliding window shift | T17: 9<br>T18: 6<br>T5: 18 |
| L2 Regularization | Backpropagation penalizes large RNN weight sizes | 0.00001 |
| Dropout | Probability of RNN units being excluded from the network during training | 0.6<br>T5: 0.5 |
| Input layer # Units | Number of units in each session specific input layer | T17: 512<br>T18: 768<br>T5: 192 |
| Input layer Dropout | Probability of input layer units being excluded from the network during training | 0.25 |
| Input activation | Input layer activation function | softsign |
| White noise SD | Standard deviation of Gaussian (zero mean) noise added to neural features (data augmentation) | 1.5 |
| Constant offset SD | Standard deviation of Gaussian (zero mean) noise added to all timesteps of neural features (data augmentation) | 0.22<br>T5: 0.1 |
| Smooth Kernel SD | Standard deviation of temporal Gaussian smoothing kernel applied to neural features | 2<br>T5: 3 |
| Batch Size | Number of sentences used to train the RNN model per step (all taken from same session in a single batch) | 64 |
| Training batches | Number of training batches (training ended early if error rate did not improve for 500 iterations) | 2,000 to 10,000 |
| Learning rate | Linearly decaying learning rate | 0.02 to 0 |
| $\beta_1$ | The exponential decay rate for the 1st moment estimates for ADAM optimizer | 0.9 |
| $\beta_2$ | The exponential decay rate for the 2nd moment estimates for ADAM optimizer | 0.999 |
| $\epsilon$ | Constant for numerical stability for ADAM optimizer | 0.1 |

**Supplemental Table 2:** 5-gram Language model parameters.

| Parameter Name | Parameter Description | Value |
| --- | --- | --- |
| Max Active | Maximum active states during beam search | 7000 |
| Min Active | Minimum active states during beam search | 200 |
| Beam | Beam length | 17 |
| Lattice Beam | Desired search through RNN logit lattice | 8 |
| Acoustic Scale | Weighting of Language model compared to RNN logit probabilities, lower value weights RNN more | 0.23<br>T5: 1.0 |
| CTC blank skip threshold | RNN logits with a blank probability exceeding this threshold are skipped during inference | 1.0 |
| Length Penalty | Negative weighting of sequence length | 0.0 |
| nBest | Number of sentences to infer in order of likelihood | 100 |
| Blank Penalty | Negative weighting applied to blank class | 4<br>T5: 1 |
| Rescore | Boolean determining G.fst rescoring of nBest inferred sentences | True |

**Supplemental Table 3:** T17 Session Data.

| **Trial Day** | **Tasks** | **Number of Sentences** |
| --- | --- | --- |
| 55 | Isolated Finger Movements + Calibration Sentences | 100 +<br>16 repetitions of each isolated finger movement condition |
| 95 | Calibration Sentences + Sentence Copy | 140 |
| 96 | Calibration Sentences + Sentence Copy + Free Typing | 122 |
| 97 | Calibration Sentences + Sentence Copy + Free Typing | 99 |
| 105 | Calibration Sentences | 40 |
| 131 | Calibration Sentences + Free Typing | 100 |
| 132 | Calibration Sentences + Free Typing | 40 |
| 144 | Calibration Sentences + Free Typing | 75 |
|  |  | **Total**<br>716 |

**Supplemental Table 4:** T18 Session Data.

| **Trial Day** | **Tasks** | **Number of Sentences** |
| --- | --- | --- |
| 43 | Isolated Finger Movements | 0 +<br>16 repetitions of each isolated finger movement condition |
| 44 | Isolated Finger Movements + Calibration Sentences | 60 +<br>4 repetitions of each isolated finger movement condition |
| 45 | Calibration Sentences + Sentence Copy + Free Typing | 78 |
| 85 | Calibration Sentences + Sentence Copy | 110 |
| 86 | Calibration Sentences + Sentence Copy | 130 |
| 92 | Calibration Sentences + Sentence Copy | 110 |
| 93 | Calibration Sentences + Sentence Copy | 130 |
| 106 | Calibration Sentences + Sentence Copy + Free Typing | 140 |
| 107 | Calibration Sentences + Sentence Copy | 160 |
|  |  | **Total**<br>918 |

